# A comprehensive evaluation methodology for the publicly accessible AI services for medical diagnostics

**DOI:** 10.1101/2021.07.23.21260626

**Authors:** Sergey P. Morozov, Victor A. Gombolevskiy, Ivan A. Blokhin, Serafim S. Semenov, Tatiana A. Logunova, Anna E. Andreychenko

**Affiliations:** Research and Practical Clinical Center for Diagnostics and Telemedicine Technologies of the Moscow Healthcare Department. 24 Petrovka Str., bldg. 1, Moscow, 127051, Russia

**Keywords:** AI (Artificial Intelligence), Teleradiology, Internet-Based Intervention, Methodology, Healthcare Quality Assurance, SARS CoV 2 Infection, Multidetector Computed Tomography

## Abstract

Online AI in telemedicine for radiology became widely available and valuable in the pandemic, particularly for chest CT analysis. On the other hand, the potentially harmful consequences of such services’ inappropriate usage cannot be neglected. Thus, a suitable methodology for quality assurance and quality control has to be established.

A study’s purpose was to develop and test an original methodology for a complex evaluation of open-access AI services in teleradiology. The approach included assessing the user experience, accessibility, safety, and diagnostic accuracy on the independent reference dataset. The methodology was applied to assess seven AI services for the detection of COVID-19 on a CT scan. A comparative analysis of this assessment is presented in this work. The analysis allowed us to draw conclusions about AI services’ quality and their value for different users – patients, physicians, and healthcare data scientists. The originality of the findings, timeliness, and interdisciplinary approach make this quality assurance methodology of particular interest for further application and spreading.

**Highlights:**

- The availability of diagnostic procedures increases each year with a corresponding increase of low-impact workload.
- During pandemics, medical AI services have become valuable in reducing the workload on healthcare professionals.
- Quality assurance for AI in healthcare requires an interdisciplinary approach - medicine, IT, and data science collaboration.
- Suggested methodology of AI quality assurance suits different target users groups, such as the general public, physicians, and data scientists.

## Introduction

In 2020, the COVID-19 pandemic spread rapidly worldwide, involving more than 120 million patients and more than 2.6 million deaths (Accessed 15 March 2021). During the COVID-19 pandemic, information technologies, especially telemedicine, have been vital worldwide, allowing patients and clinicians to communicate without contracting the infection from each other. AI services have become a full-fledged diagnostic tool in healthcare. At the same time, telehealth and AI vendors see ‘explosive’ spikes in demand, which leads to rapid AI development and implementation. As a result, there was a boost in AI-based solutions developed for COVID-19 diagnosis, especially in radiology for automated chest CT scan analysis^1-3^.

A laboratory verification (reverse transcription-polymerase chain reaction and enzyme-linked immunosorbent assay) is considered the gold standard for verifying COVID-19 infection. However, when comparing the speed of reporting and obtaining results, radiology overtakes laboratory methods. A recent study demonstrated higher sensitivity of chest CT in detecting lung involvement than the laboratory test results in cases with negative or false-negative nasopharyngeal assays^4^. Also, laboratory verification methods are associated with many requirements, such as correct material sampling, proper storage, transportation, and analysis. Non-compliance can lead to the as low sensitivity of the method as 10%^5^.

All mentioned above led to a worldwide shift in the radiology workflow^6^ with the increase in chest CT scans up to 250% during pandemic peaks accompanied by a shortage of cardiothoracic radiologists and decreased usage of other modalities and anatomical areas’ examinations^7,8^. Chest imaging in COVID-19 may be based on the recommendations of the Fleischner Society^9^. The World Health Organization recommendations divide patients into those not recommended for imaging and those for whom these studies are reasonable^10^. Faster results than molecular tests (51%) and easy access (39%) are the main reasons for using diagnostic radiology during the COVID-19 pandemic^11^. However, radiology’s efficacy and clinical value can be decreased due to the limited human resources leading to a 16.6% increase in medical errors under a requirement for the report turn-around-time reduction up to 50%^12^. In this case, the application of CAD systems and AI technologies can be beneficial for healthcare to assist radiologists in preserving high-quality reporting under the extreme conditions of a massive amount of chest CT studies as the additional diagnostic test in the COVID-19 pandemic. Simultaneously, AI technologies are developing rapidly, and many developers create services that offer an automatic AI-based CT analysis to estimate COVID-19 probability. Access to these services is open to a professional AI developer community, medical professionals, and the general public (i.e., patients).

However, no certification procedure exists for such services to indicate their quality and set guidelines on their application. Given that open online AI in telemedicine is in high demand these days and evaluation procedures are limited and can mask vulnerabilities of services^13^ and potential harmful effect on individual health, there is a need for developing a methodology and guidelines for comparative analysis and quality assurance of such services. It could lead to new standards and terms of development and using such open-access digital health services to improve management in healthcare. Therefore, this study aimed to develop and test a methodology of multiparametric AI-based telemedicine service evaluation. The interdisciplinary methodology was applied and optimized to test seven AI services for radiological diagnosis of COVID-19 during a pandemic.

## Material and methods

This study is a comparative retrospective analysis performed according to STARD criteria, centered toward the diagnostic accuracy evaluation of AI services. The independent ethics committee of the professional community of the MRS RSRR (Moscow Regional Society of the Russian Society of Radiology) was informed, and patient consent was obtained.

This study consists of two main stages: a service evaluation with the developed methodology and comparison analysis based on the evaluation results. The proposed evaluation methodology was created to test the usability, functionality, and safety of AI services on the one hand and on the other hand to assess their diagnostic accuracy on the independent reference dataset.

### Evaluation methodology

Evaluation methodology consists of assessing the usability and functionality of AI services from a general and professional user perspective and determining the services’ diagnostic accuracy on the reference dataset (Table 1).

**Table 1.** Stages of the developed methodology.

| **Methodology stage** | **Purpose** | **Methods** | **Results** |
| --- | --- | --- | --- |
| **Usability and functionality evaluation** | - to assess ease-of-use of the services from a point of view of medical professionals and general users<br>- to assess a satisfactory of target audiences with provided functions | - development of a unified questionnaire for evaluation of usability and functionality criteria<br>- a score table based on the questionnaire; | a resulting score for usability and functionality |
| **Diagnostic accuracy evaluation** | - to evaluate diagnostic performance for COVID-19 detection and assessment | ROC-analysis for classification and segmentation tasks (if available) for COVID-19 chest CT findings | sensitivity, specificity, AUC (CI 95%) |

According to potential users of AI services, the functionality options can be divided into three groups: for data scientists, who collect information and test systems; for patients, who would like to have a second opinion; for clinicians (not radiologists), who need to have results to provide treatment. A dedicated questionnaire was developed to assess the services’ user interface and functionality (Table 2). The questionnaire consists of seven subsections. Each subsection targets a particular aspect of the user interaction with the service.

**Table 2.** The developed questionnaire with relevant questions for each target user group that was utilized on the usability and functionality evaluation stage. Prioritized options for each group of users are marked in colors.

|  |  |  | Required options for user groups |  |  |
| --- | --- | --- | --- | --- | --- |
|  |  |  | Data scientists | Patients | Physicians |
| № | Criteria | Points | Max points for a group |  |  |
| 1 | Usability and functionality (15 points) |  |  |  |  |
| 1.1 | One-page website | 1 |  | 1 | 1 |
| 1.2 | Easily understandable information | 1 |  | 1 | 1 |
| 1.3 | Full lung volume analysis | 4 |  | 4 | 4 |
| 1.4 | Mobile access | 1 |  | 1 | 1 |
| 1.5 | Presence of infographics | 1 |  | 1 | 1 |
| 1.6 | Multi-language or English interface | 1 | 1 | 1 | 1 |
| 1.7 | Personal ID for the uploaded study to access results | 1 |  | 1 | 1 |
| 1.8 | Error notification during upload/analysis/returning results | 1 |  | 1 | 1 |
| 1.9 | Progress bar during study upload | 1 |  | 1 | 1 |
| 1.10 | Available on-line display of the results after processing | 1 |  | 1 | 1 |
| 1.11 | Minimum number of clicks to begin study processing (best point)<br>1-5 clicks - 2 points<br>5-10 clicks - 1 point<br>More than 10 clicks - 0 points | 2 |  | 2 | 2 |
| 2 | User instructions (2 points) |  |  |  |  |
| 2.1 | Available text and infographic instructions | 1 |  | 1 | 1 |
| 2.2 | Available video instructions (including links to Internet channels and social networks) | 1 |  | 1 | 1 |
| 3 | Available evidence-based information about the AI-algorithm (12 points) |  |  |  |  |
| 3.1 | Presence of contradictions (or limitations) | 2 | 2 |  | 2 |
| 3.2 | Information about ROC AUC | 1 | 1 |  | 1 |
| 3.3 | Sensitivity | 1 | 1 |  | 1 |
| 3.4 | Specificity | 1 | 1 |  | 1 |
| 3.5 | Information on the expected results (triage, classification and quantitative assessment) | 1 | 1 |  | 1 |
| 3.6 | Information about used clinical classifications and guidelines | 1 | 1 |  | 1 |
| 3.7 | Available scientific publications about AI service (best point)<br>Q3-Q4 scientific journals - 1 point<br>Q1-Q2 scientific journals - 2 points<br>No publications - 0 points | 2 | 2 |  | 2 |
| 3.8 | Available links to scientific publications in the service description | 1 | 1 |  | 1 |
| 3.9 | Certification (best point)<br>Available certification - 2 points<br>Warning about a lack of certification - 1 point<br>No warning about a lack of certification - 0 points | 2 | 2 |  | 2 |
| 4 | Convenient and safe study upload (3 points) |  |  |  |  |
| 4.1 | Uploading without additional conversion | 1 |  | 1 | 1 |
| 4.2 | Available information/warning about anonymization | 1 |  | 1 | 1 |
| 4.3 | Available anonymizing system | 1 |  | 1 | 1 |
| 5 | AI-service stability and speed (5 points) |  |  |  |  |
| 5.1 | Upload time (best point)<br>Less than 3 min - 2 points<br>More than 3 min - 0 points | 2 |  | 2 | 2 |
| 5.2 | Time to result after upload (best point)<br>less than 10 min – 2 points<br>more than 10 min – 1 point | 2 |  | 2 | 2 |
| 5.3 | Errors in processing (best point)<br>3% of studies or less - 1 point<br>more than 3% of studies - 0 points | 1 |  | 1 | 1 |
| 6 | Clearness of results for target audiences (5 points) |  |  |  |  |
| 6.1 | Separate only text report | 1 | 1 | 1 | 1 |
| 6.2 | Separate report containing single images with lesion markings | 2 | 2 | 2 | 2 |
| 6.3 | Modified CT series | 2 | 2 |  | 2 |
| 7 | Further actions after receiving the AI-results (3 points) |  |  |  |  |
| 7.1 | Patient management recommendations | 1 |  | 1 | 1 |
| 7.2 | Radiologist consultation provided on demand | 2 |  | 2 | 2 |
|  | **Total** | **45** | **18** | **31** | **45** |

1. Usability of services’ interface and functionality. It reflects how user-friendly is the service’ interface and the number of clicks to achieve a result — 15 points in total (33% of the final score).
2. Clearness of instructions and guidelines. Assessing the presence and quality of text or video guides or other illustrative materials explaining the service’s operation process – 2 points in total (4% of the final score).
3. Available evidence-based information about the AI model. Assessment of the provided specific information proving the model’s reliability and increasing user’s trust (possible contradictions, accuracy levels, validation results, information about available functions and expected results, links to scientific publications that contain validation results of the AI-model) – 12 points in total (27% of the final score).
4. Convenient and safe data uploading. The study is anonymized and secured by the internal service; if it cannot be anonymized, a warning appears. Convenience means no need to convert CT data from DICOM into any other format – 3 points in total (7% of the final score).
5. AI service stability is evaluated by a number of errors during data uploading or analysis. The analysis speed is compared with a relative cut-off of 10 minutes. A lengthy analysis slows a diagnostic process – 5 points in total (11% of the final score).
6. Clearness of the results implies clarity in text data and visualization. Whereas ordinary users can be satisfied with just a text conclusion about the presence or absence of COVID-19 findings, medical professionals or data scientists need more detailed information about a volume of affected areas and the service’s specifications and accuracy – 5 points in total (11% of the final score).
7. Follow-up and recommendations on the results. Patient care management with a provision of recommendations and a consultation by a radiologist on-demand is helpful for both patients and non-radiology clinicians. If a patient uses an AI service, these recommendations are necessary in every case of pathological findings, even though they are not related to COVID-19. Data scientists can neglect these recommendations in the field of research but can be taken into account for the development purpose – 3 points in total (7% of the final score).

### Statistical analysis

The evaluated AI services provided quantitative data results: a probability of COVID-19 was presented as a value from 0 to 1 or as a severity score (Thirona); a volume of the affected lung parenchyma was presented in a percentage of the total lung volume and expressed as a value from 0 to 1. If a service provided a percentage of the affected lung, it was directly used for analysis. If a service provided data in absolute values, the relative affected lung area was calculated as a ratio of the affected volume to a total lung volume. AI-models were assessed independently for each task – segmentation, and classification. ROC-analysis was performed, and AUROC values were reported with a confidence interval of 95%. Diagnostic performance was evaluated by the AUC, sensitivity, and specificity (based on the maximum Youden index) for classification and segmentation tasks of the tested AI services.

## Preparations for the evaluation

### Dataset structure

The final reference dataset included 60 anonymized chest CT studies. Forty cases were taken from a prospective registered study NCT04379531 (https://clinicaltrials.gov/ct2/show/NCT04379531). Among them, 20 positive and 20 negative COVID-19 cases were presented. Ground truth for these studies (i.e., positive or negative for COVID-19) was based on the laboratory-verified presence of coronavirus infection by RT-PCR positive test and a presence of typical COVID-19 radiological findings on CT scan. Ten different radiologists with 4-20 years of experience confirmed imaging findings. Each study was included in the dataset after reading by four randomly assigned radiologists in consensus. COVID-19 associated findings on chest CT scans included multiple peripheral ground-glass opacities, lung consolidations, and a “reverse halo” sign.

In order to test the AI model’s ability to differentiate the most socially significant lung diseases detected on chest CT scan, we randomly selected and included in our research 20 cases from the Lung Cancer Screening trial obtained in 2019 (before the COVID-19 pandemic)^14^. Fifteen cases contained CT studies with findings associated with non-viral pneumonia, and five cases had pathologically confirmed lung cancer. These cases were labeled as negative for COVID-19.

Ground truth for those studies was based on the assumption that there was no COVID-19 disease in the evaluated population before 2020. Two radiologists with 4 and 10 years of experience in thoracic radiology confirmed the presence of pathological findings on CT scans. Additional morphological verification was provided for studies with lung neoplasms.

Inclusion and exclusion criteria for the dataset are listed below. The structure and details of the resulting dataset are illustrated in Table 3.

**Table 3.**
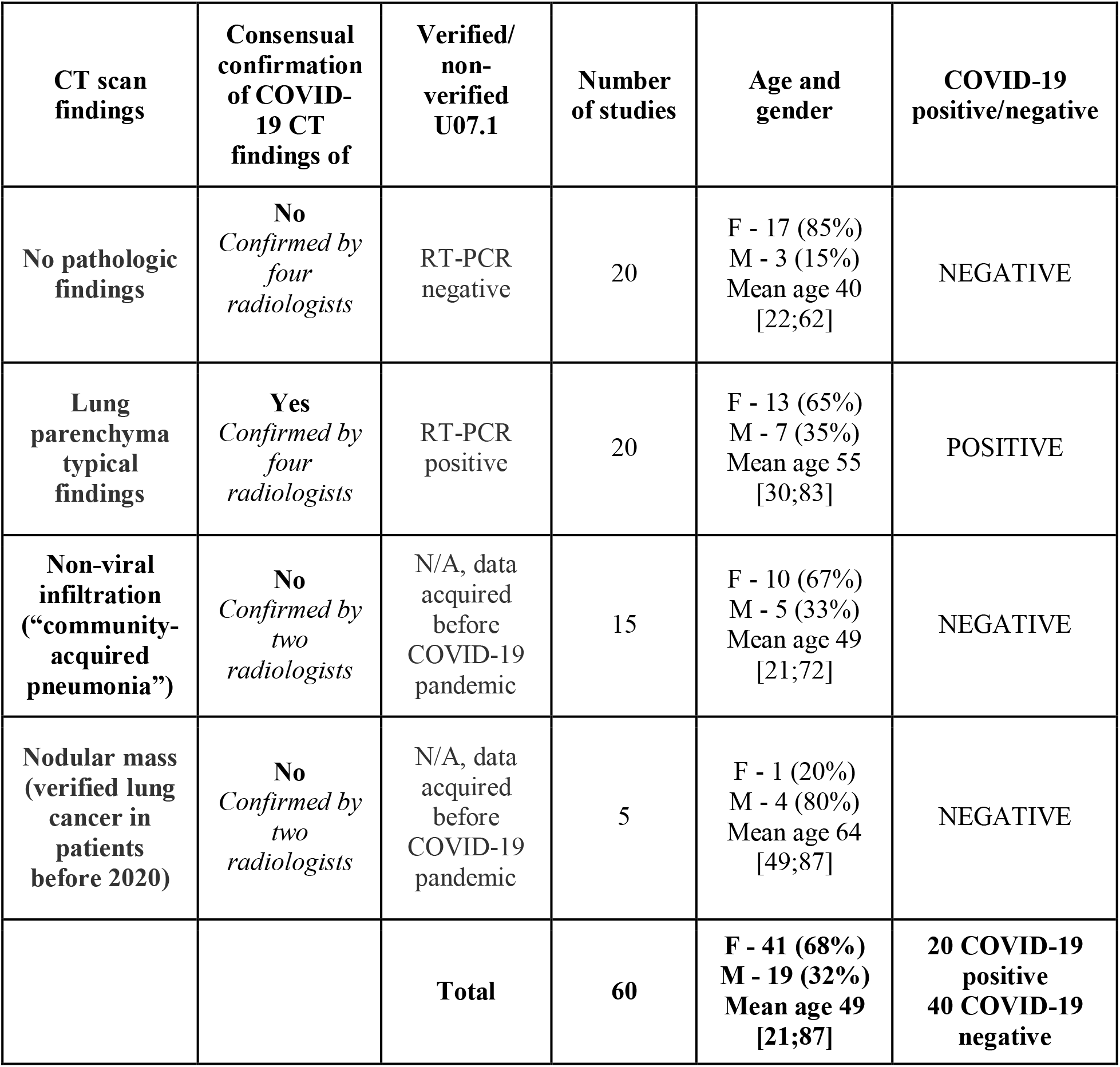
Reference dataset content

Inclusion criteria for studies in a dataset:

- only a chest region;
- inspiration breath-hold scanning;
- full chest coverage;
- spine position;
- hands at the head level.

Exclusion criteria for studies in a dataset:

- pregnancy;
- age younger than 20 years old;
- studies with incomplete chest coverage;
- studies with intravenous contrast enhancement;
- any anatomical region except a chest;
- artifacts at the lung level;
- studies of patients who underwent chest surgery.

### CT scanning protocol

All chest CT studies were obtained with a standard scanning protocol (provided by a manufacturer) on two identical 64-detector CT scanners (Toshiba Aquilion 64; Canon Medical Systems, Japan) without iterative reconstruction algorithms. The standard chest CT scan protocol had the following settings: voltage - 120 kV; rotation time - 0,5 seconds; direction - out (from legs to a head); modulation XY - on; collimation - 64*0,5 mm; helical pitch - 53,0; scanning is performed at the deep inspiration breath-hold; scan time - 6 sec on average (depending on individual constitutional features). No intravenous injection of a contrast agent was performed.

In total, each study contained two image reconstructions in an axial plane:

- reconstruction No. 1 (lungs): matrix 512*512; D-FOV - 350 mm; scan length - 300 mm (depending on individual constitutional features); reconstruction image filter - FBP QDS+; reconstruction kernel - FC51; slice thickness - 1,0 mm; increment - 1,0 mm;
- reconstruction No. 2 (soft tissues): matrix 512*512; D-FOV 350 mm; scan length - 300 mm (depending on individual constitutional features); reconstruction image filter - FBP QDS+; reconstruction kernel - FC07; slice thickness - 1,0 mm; increment - 1,0 mm.

### Additional data preparations

For AI services that were not capable of processing DICOM data, additional preparations were performed. If the AI service was intended to analyze only .jpeg images in each study (Joint Photographic Experts Group), a slice with the most visible lesion was selected and converted by free of charge viewer tool RadiAnt DICOM-Viewer Poland2016 (URL: http://www.radiantviewer.com).

### AI services search

Internet search was conducted using standard search systems and PubMed. Open-access AI services for a chest CT scan analysis were selected if the service was mentioned in the scientific publication, social media, or news or was available in a quick web search. Entry criteria for the services’ selection are listed below. In total, six services were selected for the study. Each service provides an AI model for COVID-19 analysis. Each service is available in the public domain for all Internet users.

Inclusion criteria for AI services:

- AI service with the declared capability to assess chest CT scans for COVID-19 associated findings;
- online access to AI service;
- free of charge analysis with AI models.

Exclusion criteria for AI services:

- Commercial subscription or fee-paying AI-service;
- AI service is not capable of diagnosing COVID-19.

List of selected open-access AI services for free of charge analysis of chest CT scans for COVID-19 findings and their access links:

- AiRay – https://airay.tachyhealth.com; accessed in March 2021;
- Deep Insights AI – https://labs.deep-insights.ai; accessed in March 2021;
- Medseg – http://medicalsegmentation.com/covid-19-burdenload/; accessed in September 2020;
- QUIBIM – https://imagingcovid19.quibim.com/; accessed in February 2021;
- SBERHealth – https://ai.sberhealth.ru/covid19/; accessed in October 2020;
- TelemedHUB (COVID-IRA) – http://hub.tele-med.ai; accessed in October 2020, version 1.1;
- Thirona – https://thirona.eu/cad4covid/; accessed in October 2020, version 2.0.0.

### Comparative analysis

The usability, functionality, and structure of the provided report were evaluated for each service by completing the questionnaire table (Table 2).

For comparative analysis, a total score and a score for each group were calculated with a maximum available 45 points, followed by comparing the summary results. A higher score means increased use and convenience. We analyzed the content of the provided study’s report in a similar way — if it was enough for target audiences and provided functionality required for the targeted population.

Comparison analysis of diagnostic accuracy is based on ROC-analysis results for each service in solving segmentation and classification tasks separately. The higher values of AUC correspond to higher accuracy levels. A number of false-positive and false-negative results was calculated for each service — the higher number of false reports corresponded to less valuable and reliable results.

Descriptive statistics methods with minimum and maximum values, median, first and third quartile for numerical variables, and absolute number and proportion for categorical variables were used to present the results. Nonparametric methods were used for the intergroup comparison of numerical data because the data had significant deviations from a normal distribution with a small sample size – the Kruskal-Wallis method was used to compare several groups, Mann-Whitney test was used for pairwise comparison of independent samples. The McNemar exact test was used to compare sensitivity and specificity results between different services. Given a large number of comparisons made, a p-value < 0.005 was taken as the level of statistical significance, while values of 0.005 < p < 0.05 were considered as the presence of a trend. Statistical analysis was performed using STATA14 software.

## Results

### Usability and functionality evaluation

In total, seven open access AI services for the chest CT scan evaluation for COVID-19 diagnosis were found suitable and included in the study: AiRay (https://airay.tachyhealth.com/landing/airay.html), Deep Insights AI (https://labs.deep-insights.ai), MedSeg (http://medicalsegmentation.com/covid-19-burdenload/), QUIBIM (https://imagingcovid19.quibim.com/), SBERHealth (https://ai.sberhealth.ru/covid19/), TelemedHUB (http://hub.tele-med.ai), Thirona (https://thirona.eu/cad4covid/).

The assessment of usability and functionality was made by filling in a questionnaire for each service and calculating a total score (Table 4 - compact version, Table 5 - full version).

**Table 4.**
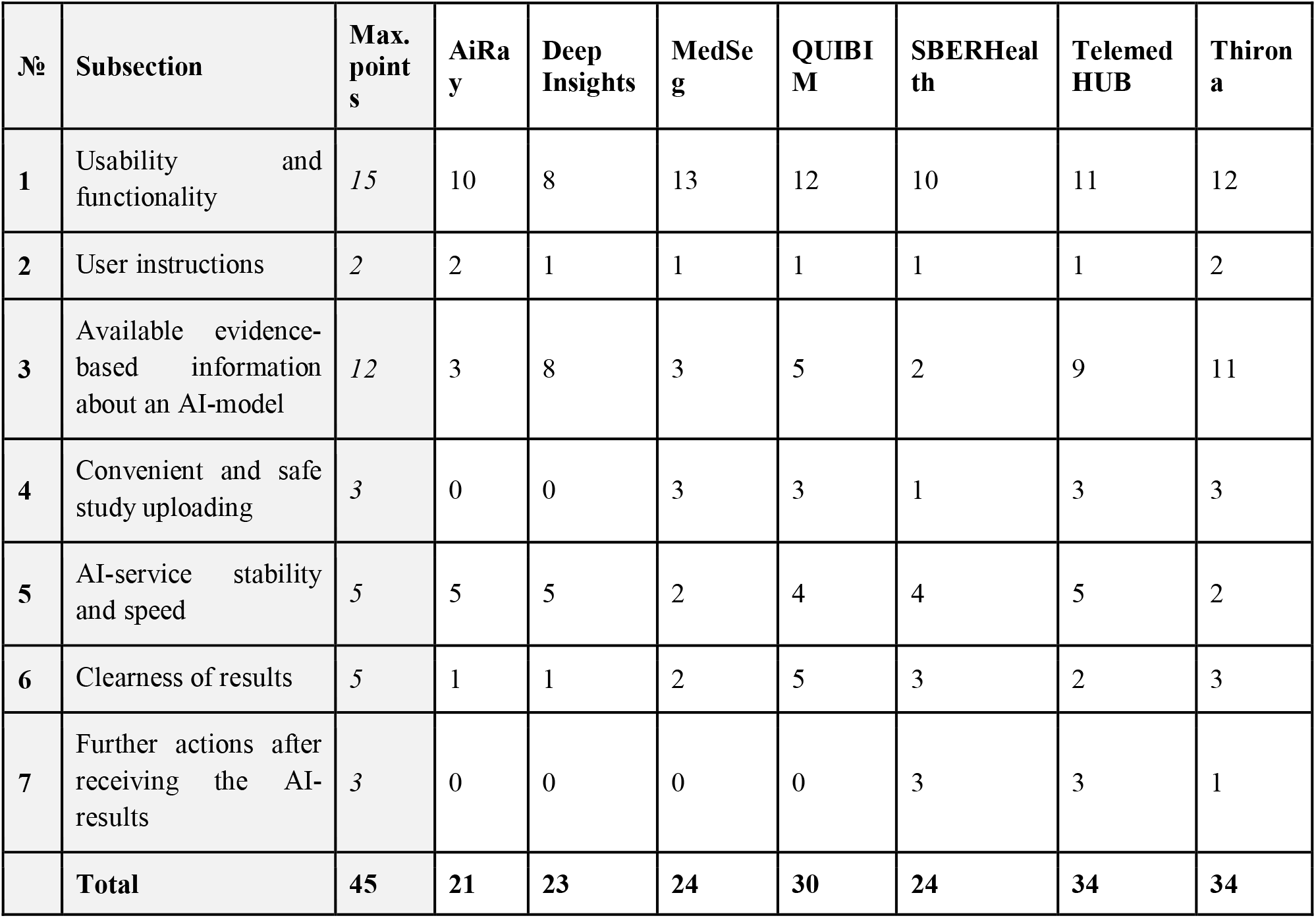
Points per subsection and total score received by each service based on the questionnaire.

**Table 5.** Completed questionnaire for usability and functionality for all tested AI-services.

| Criteria | Points | Airay | Deep Insights | Medseg | Quibim | Sber Health | Telemed HUB | Thirona |
| --- | --- | --- | --- | --- | --- | --- | --- | --- |
| Usability and functionality (15 points) |  |  |  |  |  |  |  |  |
| One-page website | 1 | 1 | 1 | 1 | 1 | 1 | 0 | 0 |
| Easily understandable information | 1 | 1 | 1 | 1 | 0 | 1 | 1 | 1 |
| Full lung volume analysis | 4 | 0 | 0 | 4 | 4 | 4 | 4 | 4 |
| Mobile access | 1 | 1 | 1 | 1 | 1 | 0 | 1 | 0 |
| Presence of infographics | 1 | 1 | 0 | 0 | 1 | 0 | 0 | 0 |
| Multi-language or English interface | 1 | 1 | 1 | 1 | 1 | 0 | 0 | 1 |
| Personal ID for the uploaded study to access results | 1 | 0 | 0 | 0 | 1 | 0 | 1 | 1 |
| Error notification during upload/analysis/returning results | 1 | 1 | 1 | 1 | 1 | 0 | 1 | 1 |
| Progress bar during study upload | 1 | 1 | 0 | 1 | 1 | 1 | 1 | 1 |
| Available on-line display of the results after processing | 1 | 1 | 1 | 1 | 1 | 1 | 0 | 1 |
| Minimum number of clicks to begin study processing (best point)<br>1-5 clicks - 2 points<br>5-10 clicks - 1 point<br>More than 10 clicks - 0 points | 2 | 2 | 2 | 2 | 0 | 2 | 2 | 2 |
| User instructions (2 points) |  |  |  |  |  |  |  |  |
| Available text and infographic instructions | 1 | 1 | 1 | 1 | 1 | 1 | 1 | 1 |
| Available video instructions (including links to Internet channels and social networks) | 1 | 1 | 0 | 0 | 0 | 0 | 0 | 1 |
| Available evidence-based information about the AI-algorithm (12 points) |  |  |  |  |  |  |  |  |
| Presence of contradictions (or limitations) | 2 | 2 | 2 | 2 | 2 | 2 | 2 | 2 |
| Information about ROC AUC | 1 | 0 | 1 | 0 | 0 | 0 | 1 | 1 |
| Sensitivity | 1 | 0 | 1 | 0 | 0 | 0 | 1 | 1 |
| Specificity | 1 | 0 | 1 | 0 | 0 | 0 | 1 | 1 |
| Information on the expected results (triage, classification and quantitative assessment) | 1 | 1 | 1 | 1 | 1 | 0 | 1 | 1 |
| Information about used clinical classifications and guidelines | 1 | 0 | 0 | 0 | 0 | 0 | 1 | 1 |
| Available scientific publications about AI service (best point)<br>Q3-Q4 scientific journals - 1 point<br>Q1-Q2 scientific journals - 2 points<br>No publications - 0 points | 2 | 0 | 0 | 0 | 0 | 0 | 0 | 2 |
| Available links to scientific publications in the service description | 1 | 0 | 1 | 0 | 0 | 0 | 1 | 0 |
| Certification (best point)<br>Available certification - 2 points<br>Warning about a lack of certification - 1 point<br>No warning about a lack of certification - 0 points | 2 | 0 | 1 | 0 | 2 | 0 | 1 | 2 |
| Convenient and safe study upload (3 points) |  |  |  |  |  |  |  |  |
| Uploading without additional conversion | 1 | 0 | 0 | 1 | 1 | 1 | 1 | 1 |
| Available information/warning about anonymization | 1 | 0 | 0 | 1 | 1 | 0 | 1 | 1 |
| Available anonymizing system | 1 | 0 | 0 | 1 | 1 | 0 | 1 | 1 |
| AI-service stability and speed (5 points) |  |  |  |  |  |  |  |  |
| Upload time (best point)<br>Less than 3 min - 2 points<br>More than 3 min - 0 points | 2 | 2 | 2 | 0 | 2 | 2 | 2 | 0 |
| Time to result after upload (best point)<br>less than 10 min – 2 points<br>more than 10 min – 1 point | 2 | 2 | 2 | 1 | 1 | 2 | 2 | 1 |
| Errors in processing (best point)<br>3% of studies or less - 1 point<br>more than 3% of studies - 0 points | 1 | 1 | 1 | 1 | 1 | 0 | 1 | 1 |
| Clearness of results for target audiences (5 points) |  |  |  |  |  |  |  |  |
| Separate only text report | 1 | 1 | 1 | 1 | 1 | 1 | 0 | 1 |
| Separate report containing single images with lesion markings | 2 | 0 | 0 | 1 | 2 | 0 | 0 | 2 |
| Modified CT series | 2 | 0 | 0 | 0 | 2 | 2 | 2 | 0 |
| Further actions after receiving the AI-results (3 points) |  |  |  |  |  |  |  |  |
| Patient management recommendations | 1 | 0 | 0 | 0 | 0 | 1 | 1 | 1 |
| Radiologist consultation provided on demand | 2 | 0 | 0 | 0 | 0 | 2 | 2 | 0 |
| **Total** | **45** | **21** | **23** | **24** | **30** | **24** | **34** | **34** |

There are two general remarks about the usability of services that are worth mentioning even though they have not been assessed directly with the developed scoring system:

- the SBERHealth included a warning that uploaded CT data have been stored in a local database for 72 hours and not transferred to third parties;
- the Thirona included an additional warning to use a widescreen monitor or connect a second one for better visualization;
- three services (SBERHealth, QUIBIM, and Deep Insights AI) displayed a warning on the main page about a limited clinical utility or research purposes only.

A probability of COVID-19 was reported by Thirona (via Severity Score), TelemedHUB, QUIBIM, AiRay, and Deep Insights AI. Five services calculated affected areas (i.e., ground-glass opacities, consolidations) and total lung volumes – Thirona, TelemedHUB, SBERHealth, QUIBIM, MedSeg.

The examples of AI service reports are presented in figure 1. Five out of seven reports contained a clear warning about the limited clinical applicability. The most detailed report was provided by QUIBIM (fig. 1d). It included tables with quantitative data about each lung’s affected area and a probability of COVID-19 or any other infectious disease. The most concise report was received from the MedSeg service. It contained a table with quantitative data about an affected area volume of the total lung volume (absolute values and percentages). Two services (AiRay (fig. 1a) and Deep Insights AI (fig. 1b)) did not generate a separate report except for a short message about COVID-19 suspicion or probability. TelemedHUB provided all the results via DICOM format.

**Figure 1.**
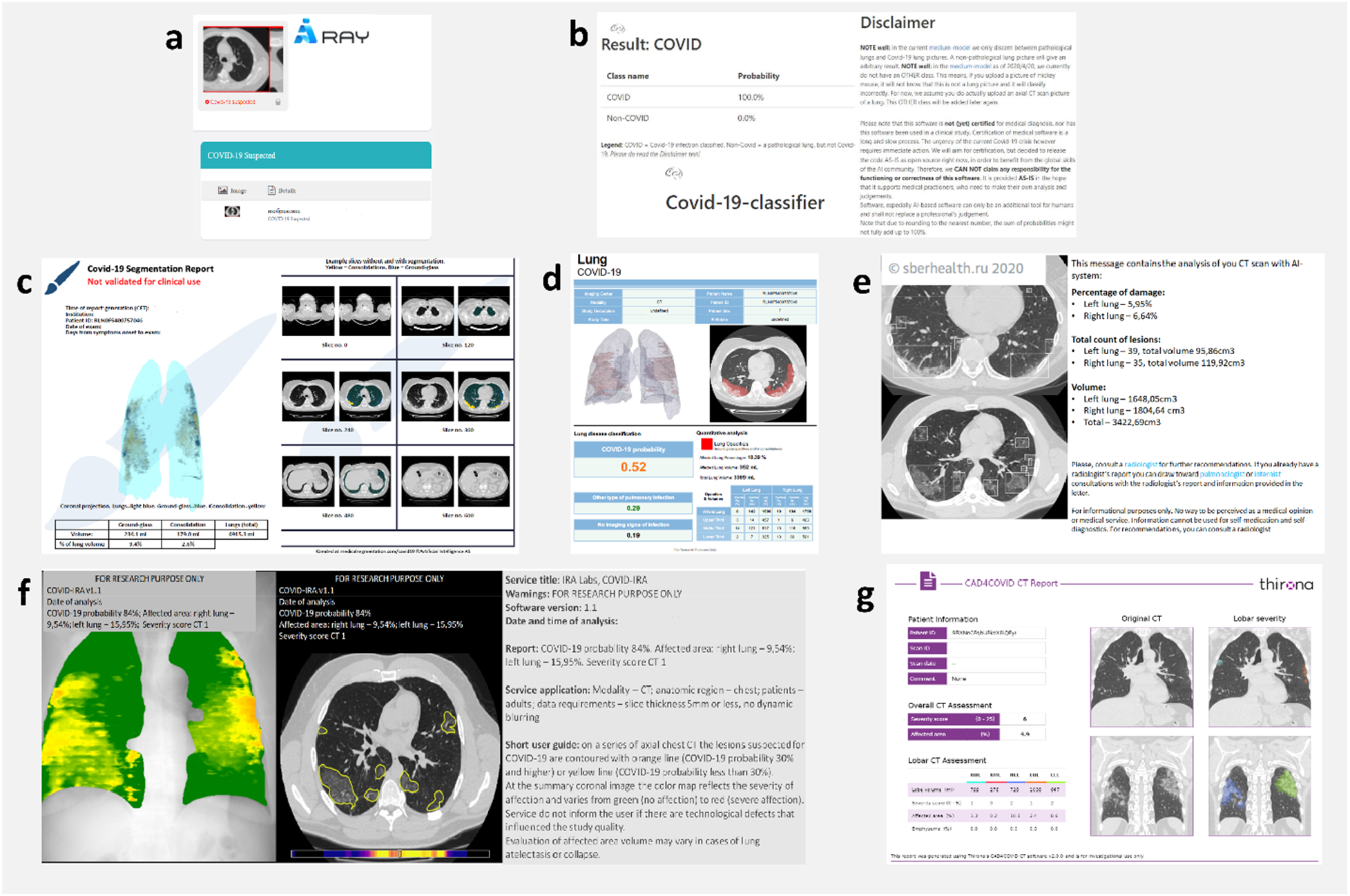
Examples of the reports provided by each service after a study analysis. a) AI Ray report contains only results for COVID-19 probability without lesion markings on the image; b) Deep Insights AI report contains only information on COVID-19 probability accompanied by the disclaimer; c) MedSeg reports COVID-19 lesions’ volume of the total lung parenchyma separately for different lesion types (ground glass-opacities and consolidations) together with 3D-rendering of lung parenchyma and several examples of axial slices with markings of the affected area; d) QUIBIM presents three probabilities (COVID-19, other infection, or no pathological changes), as well as a lesion volume and a percentage of the affected area for each lung and for the total lung parenchyma, illustrated by 3D-rendering and an example slice with markings of the affected area; e) SBERHealth reports COVID-19 lesions’ volume and a percentage of the affected area for each lung and for the total lung parenchyma, illustrations contain an additional axial CT DICOM series with markings of all lesions; f) TelemedHUB does not send a separate report file, it provides a full-sized DICOM-folder which contains text information about probability of COVID-19 and a percentage of the affected area for each lung and for the total lung parenchyma; illustrations contain a colored summary coronal slice and an additional axial CT series with markings of all lesions; g) Thirona presents results on the severity of COVID-19 disease and reports lesion volumes for each lung and for the total lung parenchyma separately for different lesion types (ground-glass opacities and emphysema); illustrations contain only two coronal reconstructions with marked lesions.

Visualization of findings on the CT images was included in five out of seven reports – either on a single axial slice (TelemedHUB, Quibim), several axial slices (Medseg, SBERhealth) or coronal slices (Thirona, TelemedHUB). 3D-rendering of COVID-19 suspected lesions in the lungs was available with MedSeg and QUIBIM. QUIBIM made illustrative examples with a single axial slice added to the report’s textual information. Stratification of ground-glass opacity and consolidation areas was available at MedSeg. Three services (TelemedHUB, SBERHealth, and QUIBIM) provided an additional separate DICOM series with marked pathological lesions.

**Usability of interface and functionality** score for all AI-services varied from moderate to high levels (from 8 to 12 points out of 15). Since an analysis of the entire study provides more reliable information, the lesser score was obtained from AI-services, which analyzed only a single image (AiRay and Deep Insights AI). Also, the smaller score was observed due to inconvenient access to results – three services did not provide online access to AI-analysis results, they were to be obtained via e-mail only and due to lack of available infographics on the website of four services (Table 2). All services provided clear user instructions on uploading a study and receiving a result in the textual form or infographics. Three services (Thirona, MedSeg, AiRay) also provided video guidance or a link to their websites.

Variations in the score concerning **available evidence-based information** about the AI model are substantial, from 3 to 11 points out of 12. The majority included descriptions of contradictions and limitations (five out of seven services), information that one can expect from analysis (five services out of seven informed the user about severity, or probability, and/or affected area’s volume). Thirona service was reported in peer-reviewed journal publications^15^, and the other services were not mentioned in scientific publications. Thirona provided open access to user manuals and specification documentation that can be helpful for medical professionals and data scientists. There was a lack of information about AI models’ diagnostic accuracy metrics and the absence of certification of four services (AiRay, MedSeg, SBERHealth, QUIBIM).

**Convenient and safe study uploading** assumes uploading of DICOM data directly without conversion to other formats and study anonymization by a provided internal tool. Two of the tested services (AiRay and Deep Insights AI) required a study conversion into .jpeg format with upload limited to a single image per time. Forced DICOM anonymization was encountered in QUIBIM – the next step of the analysis was not available without completing the anonymization procedure. One service (SBERHealth) had a warning about the absence of anonymization functionality.

**AI-service stability** was evaluated by a number of errors that occurred during uploading or analysis. The biggest number of errors was six. Two services (SBERHealth and Thirona) demonstrated six and one errors correspondingly. Five services demonstrated no errors during a study upload and analyzed all studies in the reference dataset. AI-service speed appeared to be comparable – analysis of a single study did not take more than 10 minutes, which is more than acceptable even for the emergency turnaround time for analysis of a radiological examination and a turnaround time for a notification referring clinicians^16-18^.

**Clearness of results** implies clarity in the report’s text and the presence of detailed visualization. Reported results varied from short messages about COVID-19 suspicion to complete DICOM CT series with marked findings in addition to quantitative data on severity and probability of the disease. TelemedHUB provides results in a DICOM format which requires a DICOM-viewer. The folder contains a text report illustrated by a summarized coronal slice and the entire chest CT series with lesion markings. No service had any identification of a potential target audience or any limitations for users to interpret their reports correctly.

A presence of **recommendations for a user on further actions after receiving the AI results** (3 points) was available in three services. It contained information about the necessity to consult with a radiologist or physician. Two services (SBERHealth and TelemedHUB) provided a radiologist’s consultation on-demand.

### Diagnostic accuracy assessment results performed with the reference dataset

Diagnostic accuracy was evaluated on the reference dataset containing 60 chest CT studies (20 studies with a positive result for COVID-19, 20 studies with no pathological findings, 15 studies with community-acquired pneumonia, and 5 studies with lung cancer).

Five services provided segmentation of the COVID-19 related radiological findings on chest CT scans, including ground-glass opacities, consolidations, and crazy paving. Quantitative information of the segmented areas was reported (via tables or text reports) as a percentage of the affected lung volume to a volume of the whole lung with corresponding markings on CT scans. In the case of two or more types of findings (e.g., if results were differentiated between consolidation and ground-glass opacities), their percentage was summarized to compare with the results of other services that did not distinguish the affected area types. Two services – AiRay and Deep Insights AI – did not provide results for the segmentation.

Five services provided a COVID-19 classification of the analyzed study expressed as a COVID-19 probability from 0 to 1, a severity score (0-25), or a binary decision on suspicion/non-suspicion regarding COVID-19. Thus, two separate ROC-analysis were performed for the segmentation and classification outputs (Figure 2 a,b, initial data per study can be found in Table_Raw_data.xlsx). For a confusion matrix analysis, a threshold at the maximum value of the Youden index was used. For studies with community-acquired pneumonia, lung neoplasms, and no pathological findings, if the probability of COVID-19 or value of the affected area were higher than the cut-off (maximum Youden index), the service result was labeled as a false positive. For studies with laboratory-confirmed COVID-19, if a calculated probability of COVID-19 or affected area volume was less than the cut-off (maximum Youden index), the result was labeled as a false negative.

**Figure 2.**
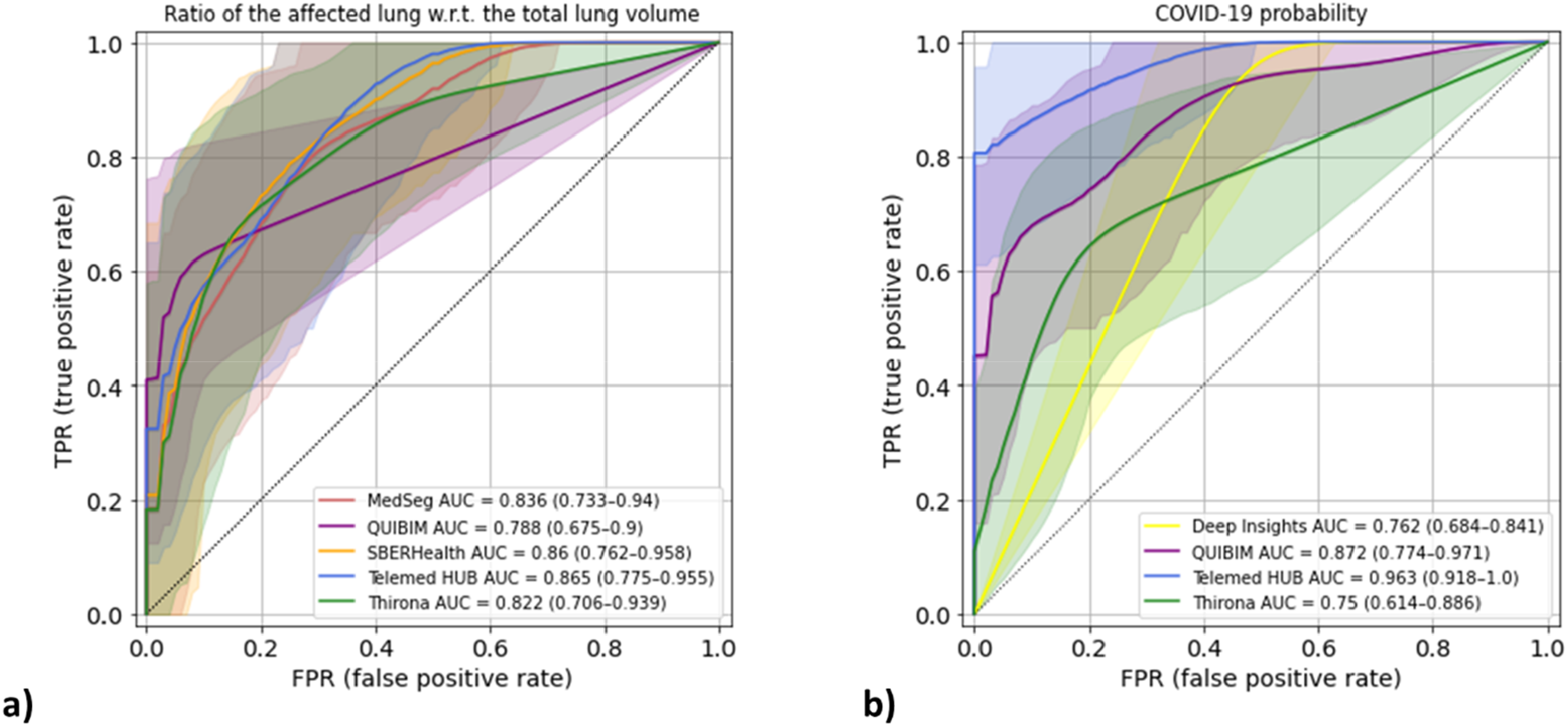
ROC curves for segmentation (a) and classification (b) tasks of the AI services for COVID-19.

The standard metrics of diagnostic performance are summarized in Table 6. Notably, an optimal operation point for the segmentation task was similar among AI models and varied from 0.4% to 0.7%.

**Table 6.** Common diagnostic accuracy metrics based on the ROC-analysis of the AI-models performance on the reference dataset

| Platform | AiRay | Deep Insights AI | MedSeg | QUIBIM |  | SBERHealth | TelemedHUB |  | Thirona |  |
| --- | --- | --- | --- | --- | --- | --- | --- | --- | --- | --- |
| Task | classification | classification | segmentation | segmentation | classification | segmentation | segmentation | classification | segmentation | classification |
| Number of tested studies (without errors) | 60 | 60 | 60 | 60 | 60 | 54 | 60 | 60 | 59 | 59 |
| True positive | 20 | 20 | 17 | 12 | 13 | 17 | 16 | 16 | 14 | 13 |
| False positive | 40 | 19 | 11 | 1 | 2 | 9 | 9 | 0 | 5 | 7 |
| False negative | 0 | 0 | 3 | 8 | 7 | 3 | 4 | 4 | 6 | 7 |
| True negative | 0 | 21 | 29 | 39 | 38 | 25 | 31 | 40 | 34 | 32 |
| Sensitivity, % | 100 | 100 | 85 | 60 | 65 | 85 | 80 | 80 | 70 | 65 |
| 95% CI | 83-100 | 83-100 | 62-97 | 36-81 | 41-85 | 62-97 | 56-94 | 56-94 | 46-88 | 41-85 |
| Specificity, % | 0 | 52 | 72 | 98 | 95 | 74 | 78 | 100 | 87 | 82 |
| 95% CI | 0-9 | 36-68 | 56-85 | 87-100 | 83-99 | 56-87 | 62-89 | 91-100 | 73-96 | 66-92 |
| Accuracy, % | 33 | 68 | 77 | 85 | 85 | 78 | 78 | 93 | 81 | 76 |
| 95% CI | 22-47 | 55-80 | 64-87 | 73-93 | 73-93 | 64-88 | 66-88 | 84-98 | 69-90 | 63-86 |
| ROC AUC, % | 50 | 76 | 84 | 79 | 87 | 86 | 86 | 96 | 82 | 75 |
| Youden index | 0.5 | 1 | 0.004 | 0.007 | 0.16 | 0.005 | 0.007 | 0.222 | 0.005 | 2.0 |

Sensitivity values of the affected lung detection varied from 60% to 85%, sensitivity values of COVID-19 probability varied from 65% to 100%. Specificity levels varied from 72% to 98% and from 52% to 100% for the affected lung detection and COVID-19 probability, correspondingly. One AI service, AiRay, demonstrated 0% specificity and was excluded from the comparative analysis.

### COVID-19 PROBABILITY COMPARISON

Four AI-services (Deep Insights, QUIBIM, TelemedHub, and Thirona) demonstrated significantly different values of a probability in COVID-19 positive and COVID-19 negative groups (p<0.001, Kruskell-Wallis test). Also, QUIBIM and Thirona AI-services demonstrated statistically significant differences in probability levels for studies with community-acquired pneumonia and lung cancer compared to studies with no pathological findings (p<0.005, Mann-Wheatney).

Comparative pairwise analysis (McNemar exact test) revealed no significant differences in sensitivity of COVID-19 probability evaluation between four AI services that provided such information (p>0.005), with a slight tendency for Deep Insights with zero false-negative results to differ from QUIBIM and Thirona, which had 7 false-negative results. TelemedHUB demonstrated intermediate sensitivity – 3 false-negative results.

Comparative pairwise analysis (McNemar exact test) revealed no significant differences in specificity levels of COVID-19 probability evaluation between QUIBIM, TelemedHUB, and Thirona (p>0.05, 32 – 40 out of 40 true negative results). Deep Insights demonstrated 21 true negative results and significantly differed from other AI services (p<=0.001). Specificity values of COVID-19 probability for Deep Insights in subgroups of community-acquired pneumonia, lung cancer, and studies with no pathological findings had a slight tendency to differ from such values for all other AI-services (0.005<p<0.05). Specificity values of COVID-19 probability for all other AI services had no significant differences (p>0.05).

### COMPARISON OF THE AFFECTED LUNG RATIO

Five AI-services demonstrated significantly different ratio values of the affected lung in COVID-19 positive and COVID-19 negative groups (p<0.001, Kruskell-Wallis test). Also, MedSeg and SBERHealth demonstrated statistically significant differences in lesion detection for studies with community-acquired pneumonia compared to studies with lung cancer and studies with no pathological findings (p<0.005, Mann-Weatney). Thirona demonstrated statistically significant differences in lesion detection for studies with community-acquired pneumonia and lung cancer compared to studies with no pathological findings (p<0.005, Mann-Wheatney).

Comparative pairwise analysis (McNemar exact test) revealed no significant differences in sensitivity of lesion detection between five AI-services that provided such information (p>0.005, 3-8 false-negative results).

Specificity values of lesion detection by QUIBIM demonstrated a slight tendency to be higher (p-values 0.008-0.04; 39 true negative results out of 40). The same tendency was revealed in a pairwise comparison of QUIBIM and SBERHealth in the subgroup of community-acquired pneumonia. Other AI services had no significant differences in the level of specificity for lesion detection.

Among the analyzed studies, there were cases when the majority of models made false predictions. However, there were no studies misinterpreted by all AI-services simultaneously. Examples of the studies that were misinterpreted by most of the services are illustrated in Figures 3, 4.

**Figure 3.**
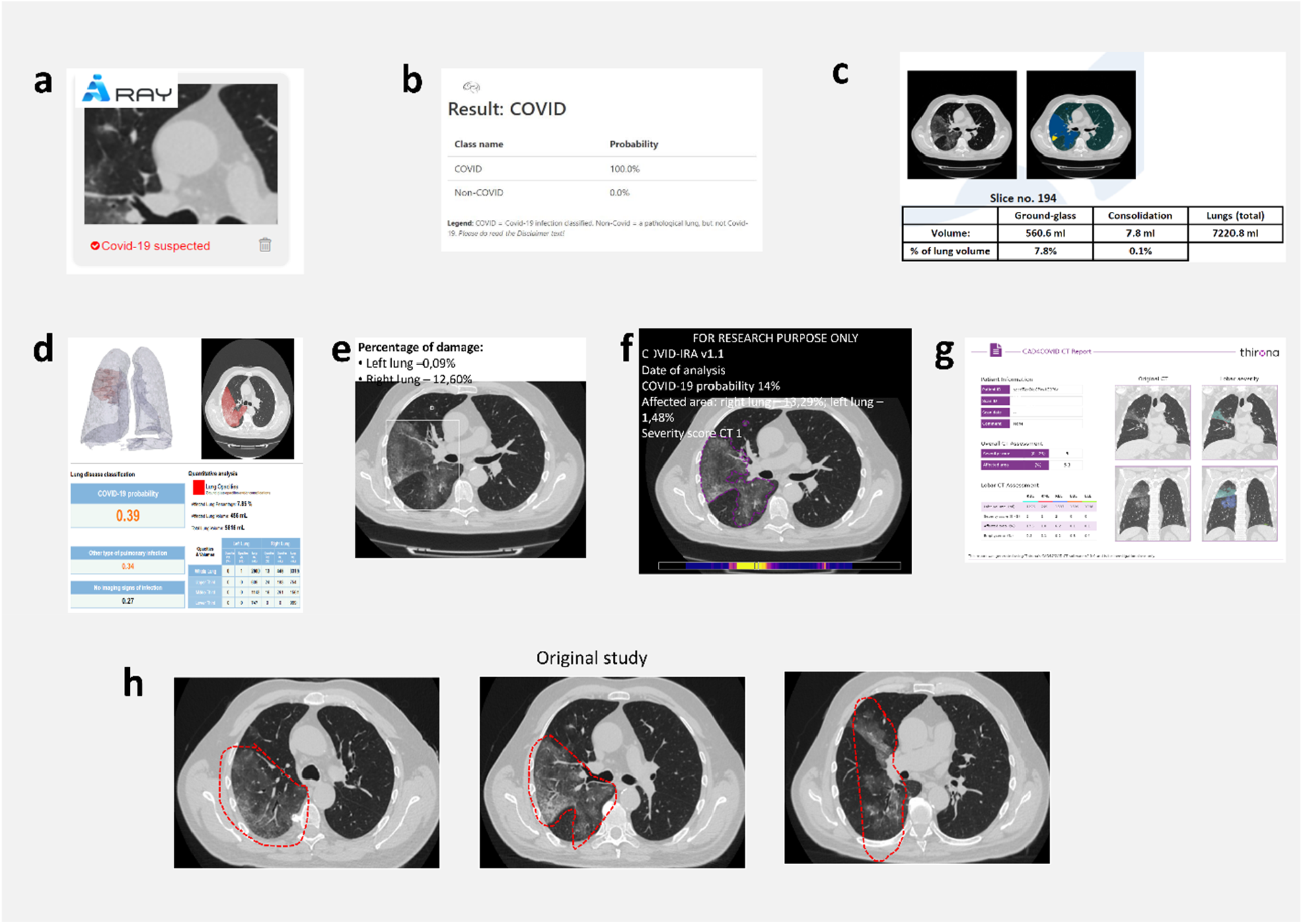
Example of a study labeled as false positive by all tested AI-services, except for the classification result by TelemedHUB. AiRay (a), Deep Insights (b), MedSeg (c), QUIBIM (d), SBERHealth (e), TelemedHUB (f), Thirona (g) determined this study as COVID-19 positive incorrectly. A correct result of the low probability of COVID-19 was provided by TelemedHUB (f). (h) Massive infiltrations in upper and lower lobes of the right lung corresponding to community-acquired pneumonia in a man in the 61-65 age group. The study was performed in 2019 before the first COVID-19 patient was registered in Russia.

**Figure 4.**
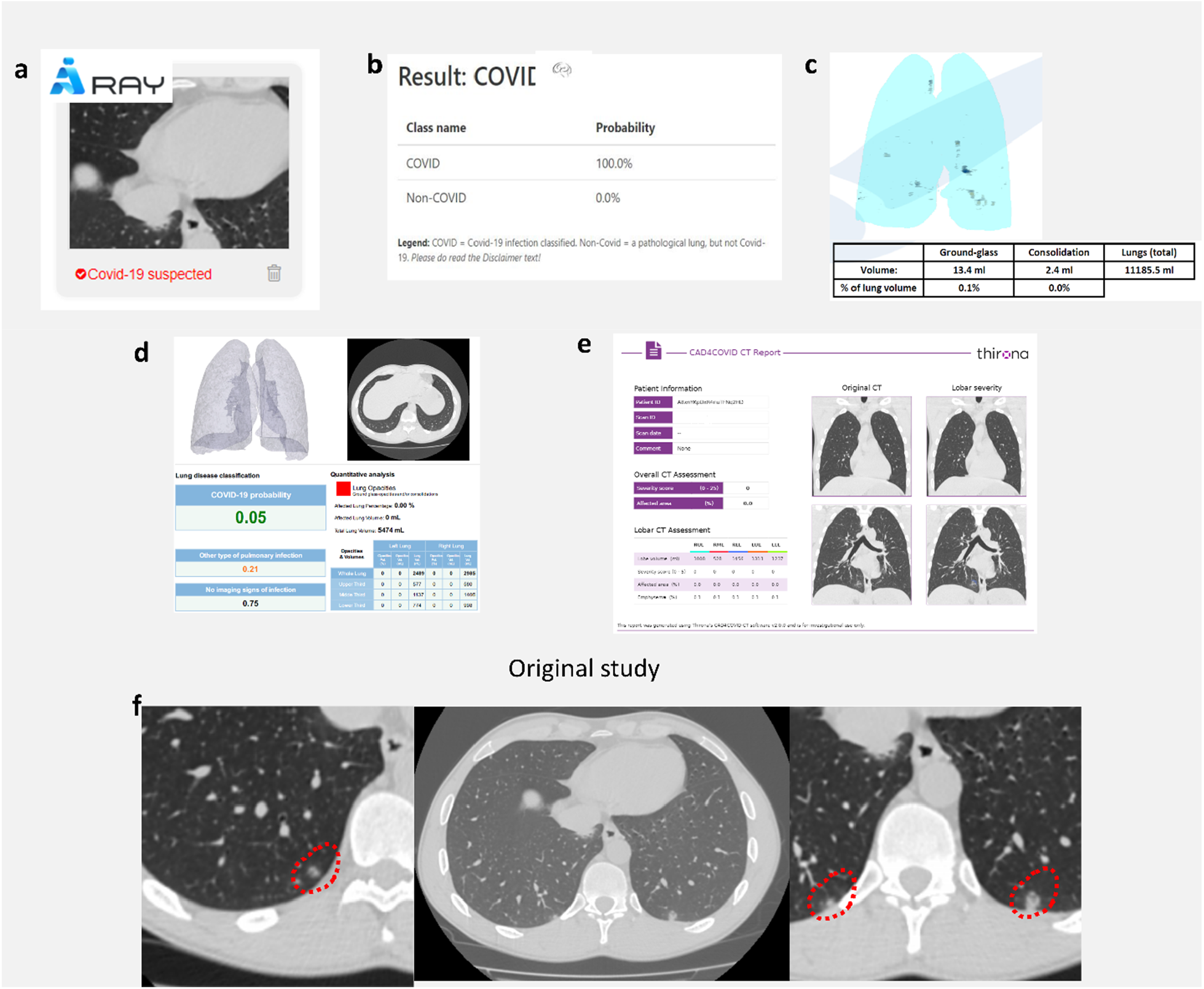
Example of study labeled as false-negative by 5 out of 7 tested AI-services. AiRay (a) and SBERHealth labeled this study as suspicious for COVID-19 correctly. Deep Insights (b), MedSeg (c), QUIBIM (d), Thirona (e), TelemedHUB^*^ determined erroneously this study as COVID-19-negative. On the original axial chest CT slice of a patient in the 26-30 age group with laboratory-confirmed coronavirus infection, there are three small lesions in lower lobes with subsolid structure and a presence of ground-glass opacity (f). ^*^Report results for SBERHealth and TelemedHUB are not presented here because the AI services had a different version online at the time of the figure creation than at the moment of the dataset analysis, so the reports were not included to this figure.

## Discussion

In this study, we used a multidisciplinary approach (medicine, IT and data science) to develop a universal methodology for evaluating the AI services that could indicate the appropriate user groups. The proposed methodology is not limited to radiology but could be used for other medical AI applications.

COVID-19 pandemic has become an excellent accelerator for developing remote radiology-related services, including education, consulting, automated diagnostic systems for COVID-19 detection and management. Simultaneously, the certification and regulations have not been matured yet, and the AI services for COVID-19 diagnosis available on the web have hardly undergone an independent quality assessment. Generally, an independent validation is necessary to separate unreliable, non-user-friendly tools that may cause harm. Nevertheless, during a pandemic, healthcare services experience a shortage of medical staff, and a need for it increases, which could be partially compensated by AI services. Different groups of users could benefit from such services: for radiologists and other medical professionals, any additional information on patients’ examinations received without delay is meaningful; for the general public and patients, it is convenient to have access to their medical information from any place and to get a professional second opinion on the study; for medical data scientists, open AI services provide valuable insights that can be used for AI research and development in healthcare. That is why many users, regardless of experience and training, continue to stay interested in such open AI services and use them despite a lack of quality assurance and certification.

For patients, high usability is important to get results quickly, without any errors, and from any place. However, the absence of explicit instructions, too many steps to get the analysis result, and a lack of data anonymization may put off the users. The availability of scientific publications on AI services and objective information about the AI-model capability and limitations may increase users’ interest and trust. It is disputable whether such information is necessary for everyday users. However, there is no doubt that scientists and medical professionals need such additional information.

While there is no clear separation of target user groups, we noticed that despite high usability levels for the public (patients), Thirona, TelemedHUB services are intended for professional usage in healthcare and research. At the same time, SBERHealth, Quibim, and MedSeg could be successfully used by general users (figure 5). QUIBIM, TelemedHUB, and Thirona services are most suitable for medical professionals’ needs. The highest usability and functionality score was obtained for services with several years’ experience in developing computer vision-based products for radiologists and scientists. It needs to be noted that a user-friendly AI report varies depending on the target audience. For example, the DICOM format of the results report in TelemedHUB does not cause any problems for trained specialists (data scientists and physicians) but can become an insuperable obstacle for general users.

**Figure 5.**
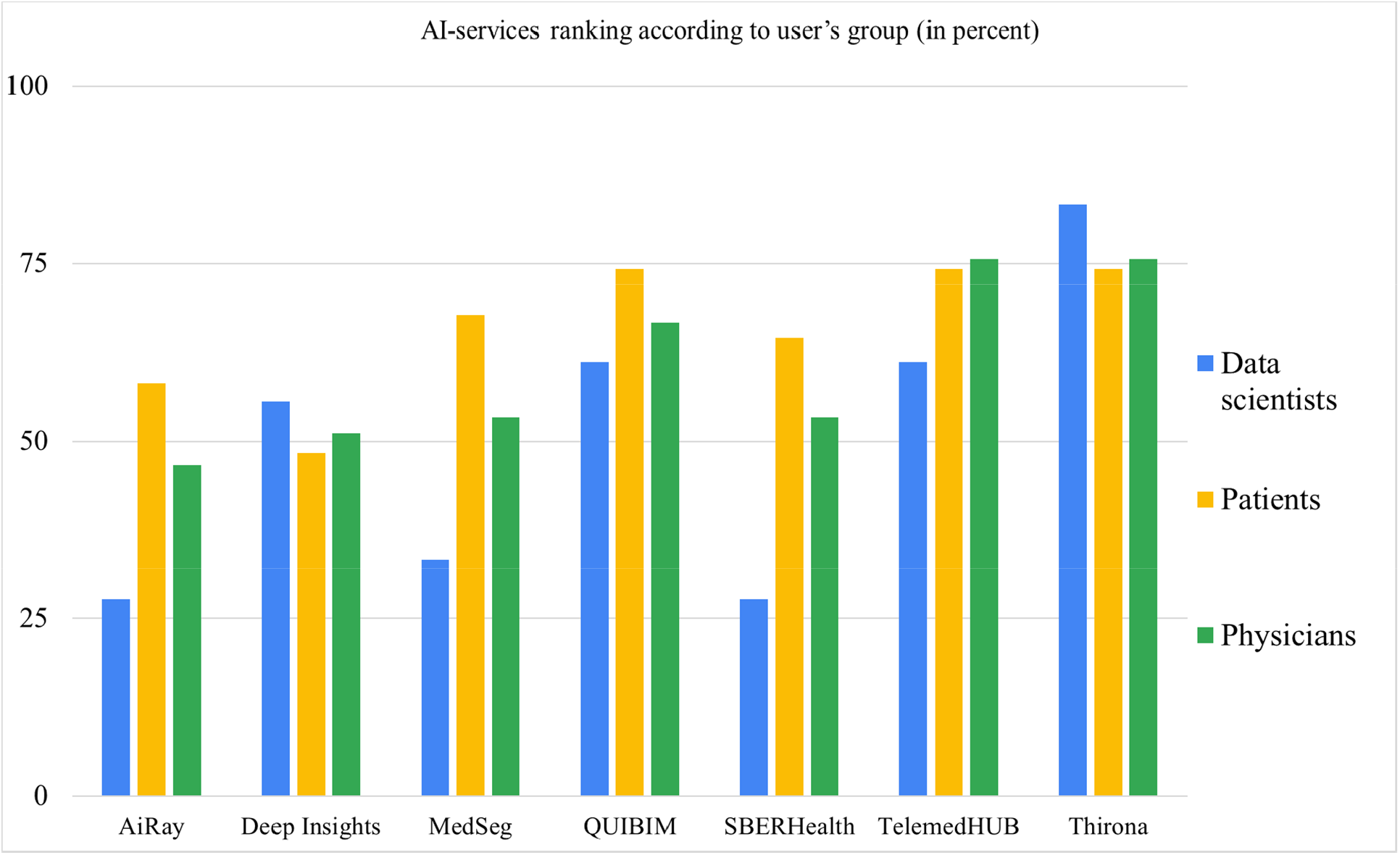
Ranking of AI-services for each user group. For medical professionals (physicians), the most conclusive results are provided by Thirona, TelemedHUB, followed by QUIBIM. For patients, results of Thirona, TelemedHUB and QUIBIM are the most conclusive; the results of SBERHealth and MedSeg are comparable and relatively high as well. For data scientists, Thirona is the most valuable, followed by QUIBIM and Telemed HUB.

The high accuracy of TelemedHUB and SBERHealth is most likely associated with the population used for model training and reference dataset since both were developed in Russia. Other AI models were trained on the different population data. Overall, our evaluation using independent data demonstrated that the services were comparable in assessing the affected area on chest CT in COVID-19, while the estimation of COVID-19 probability varied greatly – AiRay demonstrated results similar to a random guess which are not reliable. The lowest diagnostic accuracy results for COVID-19 probability evaluation by AiRay and Deep Insights AI could be explained by the limited number of analyzed slices per subject – only one axial slice converted into the image could be analyzed at once with no instructions regarding possible limitations of such an approach and a proper slice selection. It can be concluded that AI services with insufficient accuracy levels cannot be used either by medical professionals or by the general public (patients) due to a high risk of wrong decisions and errors despite the simplicity of the image upload procedure.

Since both general users and medical professionals can use open access AI services, they should operate at a high level of sensitivity. Physicians can use these services to identify patients with lung damage even if a radiologist is unavailable. Besides that, general users may easily understand the necessity of medical care. Setting a high level of specificity may be relevant to all medical professionals as it can completely change medical care. Simultaneously, AI service with high specificity and moderate sensitivity may be unwanted for general users because it may lead to self-treatment without professional medical advice.

Due to limited availability of radiologists in a pandemic, the AI services with a sensitivity reaching 100% may be of particular interest for other medical specialists as a decision support system, as well as for a non-medical public as a screening system – an early notification about a high probability of COVID-19 based on the study results can provide the opportunity to take actions at the early stage of the disease. The early detection of infected patients may save time for isolating them and starting treatment, reducing the probability of contaminating medical personal and surroundings, warning medical personnel about contact with the infected patient.

At present, the majority of offered AI services provide limited diagnostic functionality. It would be helpful for practical healthcare to provide additional information to experienced medical professionals – reports on possible outcomes, study list prioritization, possible differential diagnoses^19-21^. Despite the current limitations of AI services, we have observed a successful implementation of AI technologies during the pandemic that would stimulate their further application in healthcare on a routine basis. A flexible developers’ industry can meet demand from general users and specialists for specialized services and technologies. Thus, the impact of AI technologies in healthcare will increase among professionals and everyday users.

There are several undoubted advantages of the open-access AI services in healthcare for society:

- Disease severity assessment by AI is more robust than by physicians “by sight,” which should be considered since a shortage of radiologists in a pandemic can affect patient management.
- Reduction of the workload on medical professionals and radiologists in particular.
- Online services are more useful for patients than for practicing physicians. Medical professionals often use medical information systems with integrated services, and in most cases, are advised against external services due to personal data protection.
- In perspective, AI services can save doctors’ time, which will allow physicians to focus on challenging cases.
- Such services popularize AI by providing more clear results with an impact on the AI technology acceptance among the population – a moderate increase of trust in AI and a decrease of fear of AI.

Disadvantages of open access AI services for the non-professional community also need to be mentioned:

- A usage of non-certified AI increases the risk of unreliable results and misdiagnosis.
- The opportunity to analyze studies can lead to harmful consequences for patients if they misuse the analysis results.
- Also, getting results on their own may encourage patients to undergo unnecessary medical procedures and self-treatment.

Weighing up all advantages and disadvantages of open access AI services, we can present a list of perspectives for them:

1. Reduction of a low-impact workload for doctors; workload redistribution - simple tasks can be entrusted to an AI, and doctors may focus on more complicated cases.
2. Increasing trust in AI, reducing fear of AI.
3. The requirements and quality of open services will increase only in a competitive environment, like any commercial product.
4. New AI can capture the market faster thanks to these open services as an advertisement/promo.
5. Carrying out critical research on open services can reveal the best of them that could be safely recommended for society.
6. The market is changing with a focus on personalized and patient-centered medicine. Initially, AI models were supposed to be integrated into medical information systems and could be used only by medical professionals. With the increased amount of data for each patient, the algorithms can be personalized, increasing accuracy and reliability. Analogy: earlier, only physicians had blood pressure monitors, but nowadays, we have automatic blood pressure monitors in almost every household. However, the main conclusions and decisions will remain with doctors since AI capabilities are still limited.

## Conclusions

Quality assurance for AI in healthcare requires an interdisciplinary approach due to the complexity of human health, management systems and certain immaturity computer vision technologies/artificial intelligence technologies for healthcare. We developed a universal methodology for quality assurance of the AI services, which was successfully tested on seven radiological AI services. This methodology is applicable for different target groups of users, such as the general public (patients), physicians, and data scientists. It facilitates objective comparison of online AI services to determine their reliability, safety, and accuracy.

The availability of diagnostic procedures increases each year. A rapid increase in digital medical data has improved diagnostic accuracy and healthcare effectiveness over the last years. This tendency is going to stay for the nearest future. At the same time, the level of low-impact workload is also increasing. During a pandemic with a lack of human resources, AI services have become valuable in reducing the low-specialized workload on overburdened medical professionals via automated analysis, i.g. for tuberculosis and breast cancer screening^22,23^. However, at the moment, such services are not able to substitute medical professionals.

Such an automated approach can be spread among medical professionals and patients by developing high-quality open access AI services tailored for their user group. The purpose of publicly available AI services for the automatic analysis of radiology data is to provide its application even to untrained users. For medical professionals, it can save time to analyze more complicated and serious cases. For everyday users, automatic tools provide an opportunity to receive appropriate medical care even in lack of medical professionals and take an active part in self-care. Benefits for data scientists are in obtaining an excellent practical experience in AI development for healthcare.

Quality assurance, medical certification, and government approval are necessary for AI services and models to become full-fledged and reliable diagnostic and research tools for medical professionals, scientists, and general users.

## Supporting information

Result of the Independent Ethics Committee meeting

## Data Availability

AiRay accessed in March 2021; Deep Insights AI accessed in March 2021; Medseg accessed in September 2020; QUIBIM accessed in February 2021; SBERHealth accessed in October 2020; TelemedHUB (COVID-IRA) accessed in October 2020, version 1.1; Thirona accessed in October 2020, version 2.0.0. Dataset with CT studies is available upon request from the corresponding author.

https://airay.tachyhealth.com

https://labs.deep-insights.ai

http://medicalsegmentation.com/covid-19-burdenload

https://imagingcovid19.quibim.com

https://ai.sberhealth.ru/covid19

http://hub.tele-med.ai

https://thirona.eu/cad4covid

## Acknowledgments

We would like to express our great appreciation to several people for their contribution to this study: Y. Kirpichev for providing data analysis; N. Pavlov for assisting in the dataset formation; V. Klyashtorny, for providing statistical analysis; A. Ovsyannikov for his help in collecting the data.

## References

1. Clipper, B. The influence of the COVID-19 pandemic on technology: adoption in health care. Nurse Leader 18, 500–503 (2020).

2. Kaplan, B. Revisting Health Information Technology Ethical, Legal, and Social Issues and Evaluation: Telehealth/Telemedicine and COVID-19. International journal of medical informatics, 104239 (2020).

3. Suchá, D., van Hamersvelt, R.W., van den Hoven, A.F., de Jong, P.A. & Verkooijen, H.M. Suboptimal Quality and High Risk of Bias in Diagnostic Test Accuracy Studies on Chest Radiography and Computed Tomography in the Acute Setting of the COVID-19 Pandemic: A Systematic Review. Radiology. Cardiothoracic Imaging 2(2020).

4. Patrucco, F., et al. COVID-19 diagnosis in case of two negative nasopharyngeal swabs: association between chest CT and bronchoalveolar lavage results. Radiology 298, E152–E155 (2021).

5. Mina, M.J., Parker, R. & Larremore, D.B. Rethinking Covid-19 test sensitivity—A strategy for containment. New England Journal of Medicine 383, e120 (2020).

6. Herpe, G., et al. xCOVID-19 impact assessment on the French radiological centers: a nationwide survey. European radiology 30, 6537–6544 (2020).

7. Hashemimadani, N., Emami, Z., Janani, L. & Khamseh, M.E. Typical chest CT features can determine the severity of COVID-19: A systematic review and meta-analysis of the observational studies. Clinical Imaging.

8. Radmard, A.R., et al. A Multicenter Survey on the Trend of Chest CT Scan Utilization: Tracing the First Footsteps of COVID-19 in Iran. Archives of Iranian medicine 23, 787–793 (2020).

9. Rubin, G.D., et al. The role of chest imaging in patient management during the COVID-19 pandemic: a multinational consensus statement from the Fleischner Society. Chest 158, 106–116 (2020).

10. Akl, E.A., et al. Use of chest imaging in the diagnosis and management of COVID-19: a WHO rapid advice guide. Radiology 298, E63–E69 (2021).

11. Blažić, I., Brkljačić, B. & Frija, G. The use of imaging in COVID-19—results of a global survey by the International Society of Radiology. European radiology 31, 1185–1193 (2021).

12. Berlin, L. Faster reporting speed and interpretation errors: conjecture, evidence, and malpractice implications. Journal of the American College of Radiology 12, 894–896 (2015).

13. Wu, E., et al. How medical AI devices are evaluated: limitations and recommendations from an analysis of FDA approvals. Nature Medicine, 1–3 (2021).

14. Morozov, S.P., et al. Low-dose computed tomography in Moscow for lung cancer screening (LDCT - MLCS): baseline results. Voprosy onkologii 65, 224–233 (2019).

15. Lessmann, N., et al. Automated Assessment of COVID-19 Reporting and Data System and Chest CT Severity Scores in Patients Suspected of Having COVID-19 Using Artificial Intelligence. Radiology 298, E18–E28 (2021).

16. Do, H.M., et al. Augmented radiologist workflow improves report value and saves time: a potential model for implementation of artificial intelligence. Academic radiology 27, 96–105 (2020).

17. Perotte, R., et al. Improving emergency department flow: reducing turnaround time for emergent CT scans. in AMIA Annual Symposium Proceedings, Vol. 2018 897 (American Medical Informatics Association, 2018).

18. Towbin, A.J., et al. Practice policy and quality initiatives: decreasing variability in turnaround time for radiographic studies from the emergency department. Radiographics 33, 361–371 (2013).

19. Aggarwal, R., et al. Diagnostic accuracy of deep learning in medical imaging: a systematic review and meta-analysis. npj Digital Medicine 4, 1–23 (2021).

20. Loria, K. Putting the AI in radiology. Radiology Today 19(2018).

21. Neri, E., et al. What the radiologist should know about artificial intelligence-an ESR white paper. (2019).

22. Qure.ai. https://qure.ai April 19, 2021

23. Lunit INSIGHT. https://insight.lunit.io April 19, 2021

