## Supplementary material for "A comprehensive evaluation methodology for the publicly accessible AI services for medical diagnostics": Result of the Independent Ethics Committee meeting

### Independent Ethics Committee of Moscow Regional Office of the Russian Society of Radiologists and Radiographists

*28 Srednyaya Kalitnikovskaya St., bld. 1, Moscow;*

**Approval Number No. 2 (1-II-2020) of the meeting of IEC of RSRR MRO dated February 20, 2020**

**Present:** O.A. Agafonova, E.G. Bahteeva, A.S. Laipan, I.S. Komolov O.A. Mokienko, I.I. Nadelyaeva, L.A. Nizovtsova, O.V. Omelyanskaya, A.V. Petraikin

**Topics:** Expert review of the documents for the upcoming study titled the "Experiment on the use of innovative technologies in the field of computer vision for the analysis of medical images and further use in the healthcare system of Moscow."

**Documents submitted for review:**

1. The abstract of the study titled the "Experiment on the use of innovative technologies in the field of computer vision for the analysis of medical images and further use in the healthcare system of Moscow"
2. A cover letter to the attention of the Chairman of IEC of RSRR MRO concerning the ethical expert review of the study titled the "Experiment on the use of innovative technologies in the field of computer vision for the analysis of medical images and further use in the healthcare system of Moscow"
3. CV of the Principal Investigator
4. A patient information and informed consent form
5. A project overview.

**Resolution:** To approve the upcoming study titled the "Experiment on the use of innovative technologies in the field of computer vision for the analysis of medical images and further use in the healthcare system of Moscow."

***Due to the conflict of interest O.A. Agafonofa, the co-implementer of the project, did not take part in the vote.***

Chair of IEC of RSRR MRO: [signature]
